# Temporal stability and detection sensitivity of the dry swab-based diagnosis of SARS-CoV-2

**DOI:** 10.1101/2021.05.28.21258007

**Authors:** CG Gokulan, Uday Kiran, Santosh Kumar Kuncha, Rakesh K Mishra

**Affiliations:** CSIR-Centre for Cellular and Molecular Biology (CSIR-CCMB), Hyderabad, India; Academy of Scientific and Innovative Research (AcSIR); Buchmann Institute for Molecular Life Sciences, Goethe University Frankfurt, Germany

**Keywords:** COVID-19, Viral strains, Diagnosis, Stability, Proteinase K, Dry swab

## Abstract

The rapid spread and evolution of various strains of SARS-CoV-2, the virus responsible for COVID-19, continues to challenge the disease controlling measures globally. Alarming concern is, the number of second wave infections surpassed the first wave and the onset of severe symptoms manifesting rapidly. In this scenario, testing of maximum population in less time and minimum cost with existing diagnostic amenities is the only possible way to control the spread of the virus. The previously described RNA extraction-free methods using dry swab have been shown to be advantageous in these critical times by different studies. In this work, we show the temporal stability and performance of the dry swab viral detection method at two different temperatures. Contrived dry swabs holding serially diluted SARS-CoV-2 strains A2a and A3i at 25°C (room temperature; RT) and 4°C were subjected to direct RT-PCR and compared with standard VTM-RNA based method. The results clearly indicate that dry swab method of RNA detection is as efficient as VTM-RNA-based method in both strains, when checked for up to 72 hours. The lesser C_T_ values of dry swab samples in comparison to that of the VTM-RNA samples suggest better sensitivity of the method within 48 hours of time. The results collectively suggest that dry swab samples are stable at RT for 24 hours and the detection of SARS-CoV-2 RNA by RT-PCR do not show variance from VTM-RNA. This extraction free, direct RT-PCR method holds phenomenal standing in the present life-threatening circumstances due to SARS-CoV-2.

## Introduction

The life-threatening and repeated Coronavirus Disease 2019 (COVID-19) pandemic waves clubbed with the emergence of new strains and scarceness of the vaccines for majority of the world population, the only way out is to follow COVID-19 protocols and mass testing to stop the spread of the virus. In a year’s time the COVID-19 testing facilities has evolved to test more samples by implementing automation and involving more skilled people. Regardless of all these preparations, due to multiple waves of infection and new strains with fast spreading ability, the present testing setup is unable to test the large number of samples in a time bound manner. Along with this, there are multiple challenges such as shortage of Ribonucleic acid (RNA) isolation reagents, samples transport facilities, equipment breakdown, and limited human/financial resources (Fomsgaard & Rosenstierne, 2020). In an attempt to overcome these hurdles, multiple studies attempted to present the advantages of RNA extraction-free direct Reverse Transcription-Polymerase Chain Reaction (RT-PCR), without compromising the sensitivity of detection (Bruce et al., 2020; Kiran et al., 2020; Padgett et al., 2020; Parikh et al., 2021; Smyrlaki et al., 2020; Visseaux et al., 2021) which is being effectively implemented in few countries (ICMR, 2021; Srivatsan, Heidl, et al., 2020).

Besides the existence of promising extraction-free RT-PCR methods, there are apprehensions in implementing such methods in large scale. Those include temporal stability of the samples, detection sensitivity, other logistics, and mainly the fact that diagnostic personnel being accustomed to traditional methods. But the current circumstances demand more flexibility to implementation of extraction-free detection methods that saves considerable time, manpower, and reduce the economic burden. In this work we convincingly added more scientific dimensions to the dry swab extraction-free method for Severe Acute Respiratory Syndrome-Coronavirus-2 (SARS-CoV-2) diagnosis that we have reported earlier (Kiran et al., 2020). This work illuminates further advantages of the method and discusses the scope for commercialization.

## Materials and methods

### Viral strains, log dilutions and swab preparation

Strains representing two different clades of SARS-CoV-2, namely A2a and A3i (GISAID ID: EPI_ISL_458046; virus ID-hCoV-19/India/TG-CCMB-L1021/2020, and EPI_ISL_458075; virus ID-hCoV-19/India/TG-CCMB92 O2-P1/2020) have been reported previously and have been tested for the temporal stability (Gupta et al., 2021). A concentration of 4.55×10^6^ pfu/ml and 8×10^6^ pfu/ml viral cultures of A2a and A3i strains, respectively, were serially diluted to up to 6 log folds. The swabs were immersed in different dilutions for 10-15 seconds and then stored as either dry swab in 15 ml sterile tubes or in 3 ml VTM-containing tubes at room temperature (RT; 25±1□) and 4°C for up to 3 days. Independent swabs were sampled at an interval of 24 hours and processed as discussed below.

### Sample processing

The sample processing was performed in the BSL-3 facility of CSIR-CCMB, Hyderabad, India by following Standard Operating Procedures.

#### a) Extraction of biological material from dry swabs

A volume of 400 μl of TE buffer [Tris pH-7.4 10 mM, EDTA 0.1 mM] with 2mg/ml proteinase K was added and the samples were incubated at RT for 30 min to ensure the release of biological material (unless mentioned otherwise).

#### b) Heat Inactivation

The samples were than heated at 98° C for 6 min in a dry bath placed inside the laminar hood. The heat-treated samples were directly used as a template for RT-PCR.

### Proteinase K concentration optimisation

Nasopharyngeal swab samples from three COVID-19 positive individuals with informed consents were collected. 400 μl of TE buffer was added to the swab samples at first and the samples were let at RT for 15 minutes. Later, four 50 μl aliquots of the samples were taken and appropriate volume of 100 mg/ml proteinase K solution was added to the aliquots, respectively, to get 2 mg/ml, 1 mg/ml, 0.5 mg/ml of proteinase K. One tube was processed without proteinase K (0 mg/ml). The samples were let at RT for another 15 minutes and were processed as described above.

### RNA extraction and RT-PCR

The RNA was extracted from the VTM samples using QIAamp Viral RNA isolation kit (Qiagen, Germany) by following the manufacturer’s protocol. The RNA samples were checked for the presence of SARS-CoV-2 RNA using FDA (Food and Drug Administration, USA Government)-approved Fosun COVID-19 RT-PCR Detection Kit (Shanghai Fosun Long March Medical Science Co., Ltd, China). The kit provides primers and probes for target genes, the envelope gene (E-gene; ROX labelled), nucleocapsid gene (N-gene; JOE labelled), and open reading frame1ab (ORF1ab; FAM labelled) of SARS-CoV-2. The RT-PCR was performed as per manufacturer recommendation on QuantStudio™5. The Positive and negative controls provided in the kit were included each time in the amplification plates, and the C_T_ values were in the range of the manufacturer protocol proving to be efficient and devoid of contamination. All the samples were tested in triplicates.

### Statistical analysis

Microsoft excel was used for statistical analysis and plotting. GraphPad prism 6 was also used for plotting.

## Results

### Detection of SARS-CoV-2 RNA is independent of storage temperature till 24 hours

One of the major concerns for adopting dry swab-based diagnosis at large scale is the stability of the viral RNA during shipping and storage. Presence of intact RNA is essential for reliable diagnosis, and the integrity of RNA is dependent on multiple factors of which temperature is a crucial factor. Here, the detection of SARS-CoV-2 RNA was assessed by subjecting contrived swabs in VTM and contrived dry swabs at two temperatures for different periods of time. The results indicate that storing the contrived dry swabs at room temperature (RT; 25±1□) does not affect the detection ability for up to 24 hours. In addition, the C_T_ values of the dry swab samples and that of the RNA isolated from corresponding VTM samples were comparable when stored at RT. Similarly, samples stored at 4□ showed no differences when incubated in VTM and as dry swabs for 24 hours (Figure 1; Supplementary File 1).

**Figure 1:**
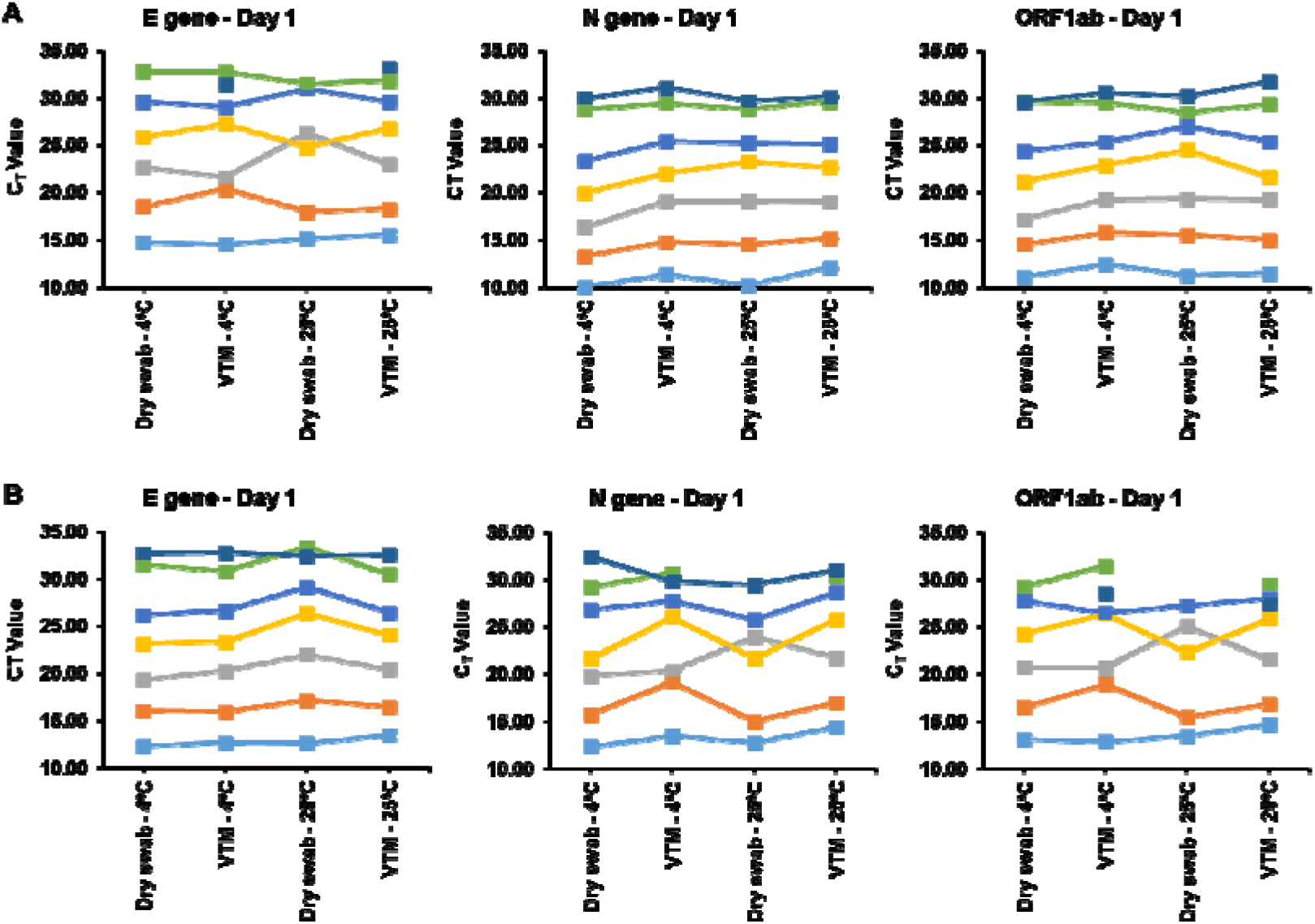
Stability of the viral RNA is unaffected by different storage conditions for up to 24 hours of storage. C_T_ values of the viral genes in (A) A2a strain and (B) A3i strain after 1 day of storage in the mentioned conditions. RT-PCR was carried out in triplicates for every sample and the average C_T_ values are plotted. Each coloured line represents one sample.

### Long-term stability of the viral RNA is temperature-dependent

While the above results provide the stability of viral RNA unperturbed up to 24 hours at RT, the long-term storage/shipping for performing genome analysis is unexplored. Therefore, we studied the effect of long-term storage of the contrived samples at different temperatures. Results indicated that storing the samples at 4□ could help in maintaining the integrity of RNA for up to 3 days irrespective of the presence or absence of VTM. The C_T_ values of the VTM samples and dry swab samples stored at 4□ were comparable for majority of the samples suggesting that storing the swabs in the absence of media does not affect the integrity of the RNA (Figure 2; Supplementary File 1). However, when stored at RT, the dry swab samples showed increased C_T_ indicating the loss of RNA copies after 2 days of storage. As a result, samples with low viral titres went undetected. Even so, samples stored in VTM at RT, and 4□ and the dry swabs stored at 4□ remained intact as suggested by the C_T_ values that are comparable to a greater extent (Figure 2).

**Figure 2:**
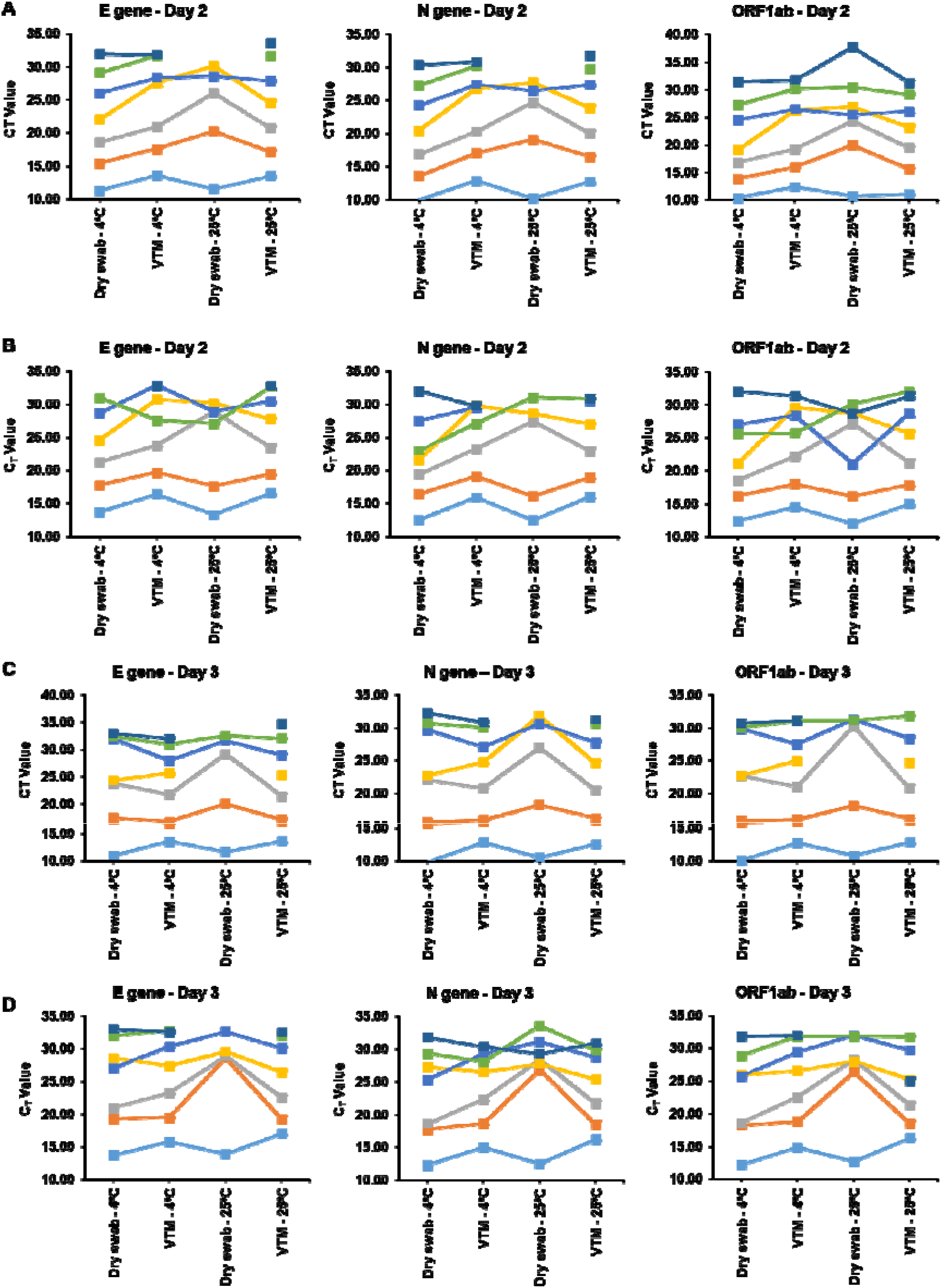
Long-term storage of dry swab samples at RT causes loss in detection of low titre samples. CT values of the viral genes in (A) A2a strain and (B) A3i strain after 2 days of storage and similarly that of (C) A2a strain and (D) A3i strain after 3 days of storage. RT-PCR was carried out in triplicates for every sample and the average CT values are plotted. Each coloured line represents one sample.

### Prolonged storage of dry swabs at 4□ is as efficient as storing in VTM

The reduced viral RNA integrity upon storage at higher temperatures in different media has been previously reported (Druce et al., 2012). To overcome this problem of reduced stability the storage of samples at lower temperatures can be of probable solution. In this regard, a comparison of the samples stored in different conditions revealed a temporal increase in the C_T_ values of the dry swab samples when stored at RT but not at 4□. The median C_T_ values of the dry swab samples stored at 4□ at all time points are comparable to that of the VTM samples (Supplementary Figure 1). Statistical correlation analyses revealed that there is a strong positive correlation between the C_T_ values of dry swab samples and VTM-RNA when stored at 4□ for 3 days and at RT for 1 day (Figure 3A; Supplementary Table 1). Moreover, the C_T_ values of the dry swab samples stored at 4□ showed strong correlation between 3 days and 1 day of storage (Figure 3B; Supplementary Table 2). Together, the data suggest that dry swab-based sample collection could be a potential alternate for sample collection in VTM given the availability of refrigeration and when the samples has to be stored or transported for a longer time. Additionally, the delta C_T_ values between dry swab and VTM-RNA confirms the validity of the above claim, wherein the dry swab samples stored at 4□ shows negative to no difference in the delta C_T_ values when compared to VTM-RNA samples (Supplementary Figure 2).

**Figure 3:**
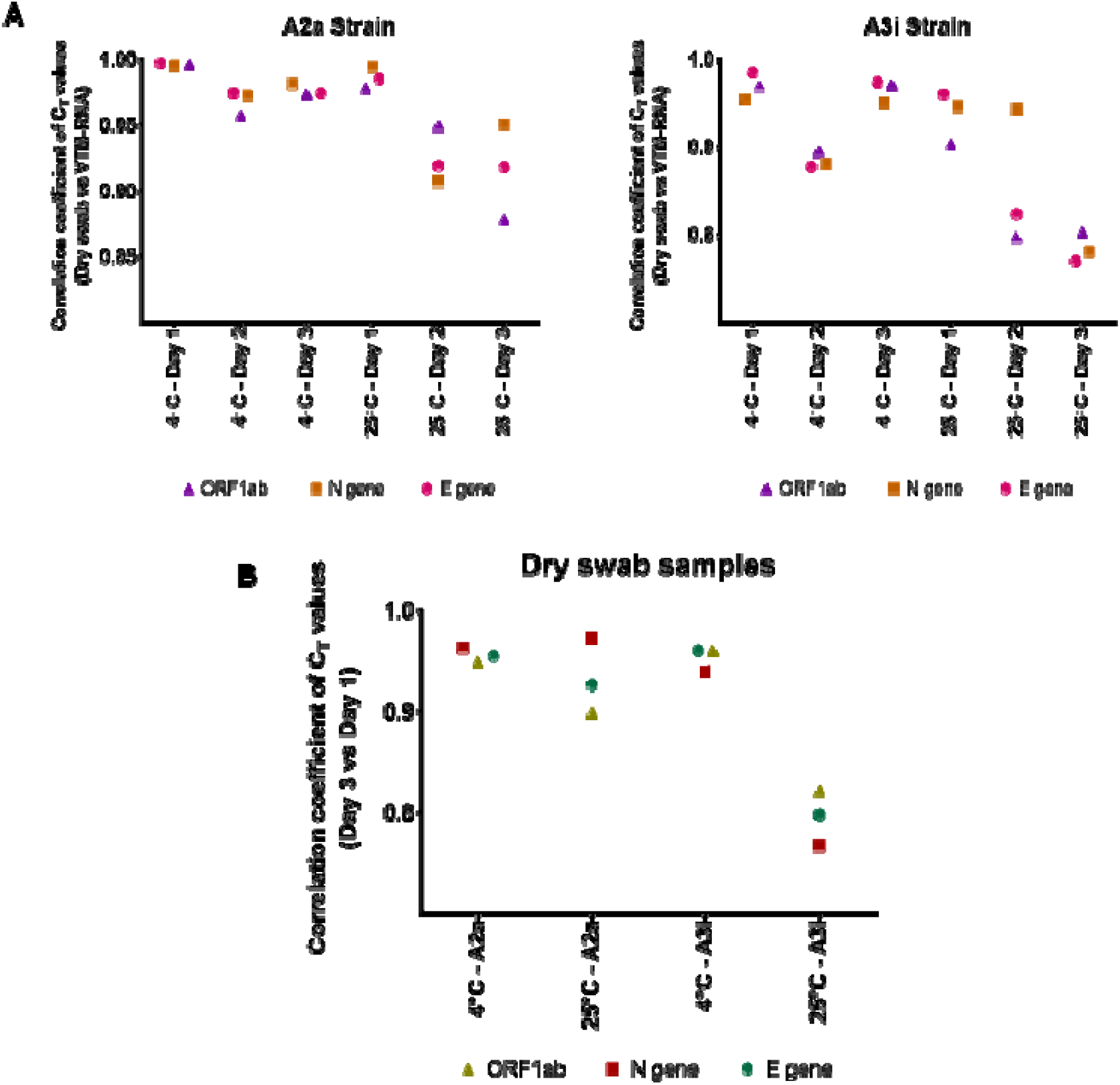
Scatter plots showing (A) the comparison of the correlation coefficients between the C_T_ values of the dry swab and VTM-RNA samples subjected to the given conditions and (B) the correlation between the C_T_ values of the dry swab samples between day 3 and day 1. RT-PCR was performed in triplicates for each sample and the average C_T_ values were used to calculate the correlation coefficient using.

### Optimization of proteinase K concentration

Use of proteinase K in RNA extraction-free methods is reported to increase the detection efficiency (Srivatsan, Han, et al., 2020). The FDA-authorized SalivaDirect method also uses proteinase K at a concentration of 2.5 mg/ml as the major ingredient for effective detection (Vogels et al., 2020). For the dry swab method using TE buffer and proteinase K, the major cost component is proteinase K and finding the minimum concentration of proteinase K could help in increasing the throughput of tests with a given amount of the enzyme thereby cutting down the per test cost. In an attempt to optimise the concentration of proteinase K required for efficient detection, we tested dry swabs from three independent COVID-19 positive nasopahryngeal samples with different viral loads at four proteinase K final concentrations. Results suggest that a final concentration of 0.5 mg/ml of proteinase K produces comparable with that of 2 mg/ml (Figure 4). In accordance with previous reports, lack of proteinase K affected the detection as suggested by increased C_T_ values when compared to samples with proteinase K (Figure 4; Supplementary Table 3).

**Figure 4:**
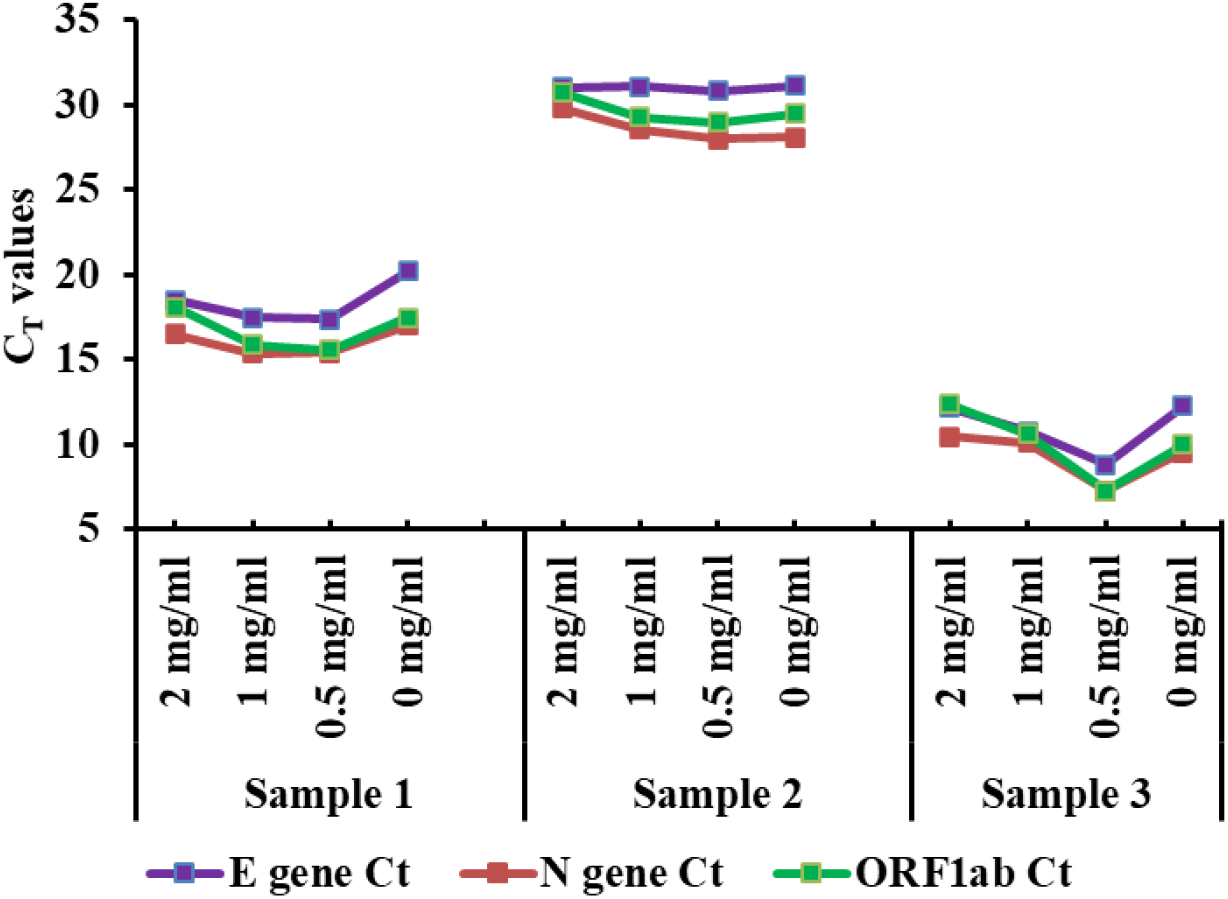
Line chart showing the effect of proteinase K concentration on the detection of SARS-CoV-2 targets in 3 independent COVID positive individuals’ samples.

## Discussion

Considering the exponential raise in the number of SARS-CoV-2 infected people and hospitalisations, it is imperative to look for alternative methods to quickly diagnose and control the disease spread. Based on the availability of resources and sample burden, diagnostic labs need to adopt methods with faster turnover time to cater the present health care demands. Here in this work, we presented an additional advantage of the dry-swab method over time consuming traditional way and highlighted the elimination of cold chain for shorter sample transport. The study on temporal stability of the two strains of SARS-CoV-2 at two different temperatures indicates that, for shorter distance transportation cold chain can be avoided and the dry swab samples with low viral load also is stable at RT for 24 hours. In case of high sample surge the swabs can be stored at 4°C for up to 3 days without compromising the detection sensitivity. The dry swab method is expected to hold similar advantages in case of other emerging variants of SARS-CoV-2 as indicated by multiple sewage surveillance and sequencing studies (Crits-Christoph et al., 2021; Martin et al., 2020). This suggests that the virus variants could be different with respect to the rate of infection and eliciting immune response, but they are generally stable for longer time in different conditions. Additionally, as a measure to cut cost incurred per test, we optimised the minimum final concentration of proteinase K that can produce reliable results. Our results indicate that a minimum final concentration of 0.5 mg/ml of proteinase K is enough to produce results comparable to that of 2 mg/ml. This evidently suggests that the throughput of the tests can be increased four times in addition to bringing down the cost by four times.

In conclusion, the dry swab method facilitates multiple advantages at the present critical time while the testing facilities are failing to cater the requirements. With the present testing facilities and resources, the dry swab method can increase the throughput of a lab by three-folds. The challenges like reagent shortage, limited human resources and high transmission rate can be handled in a better manner. The recent study on culturing virus from dry swab clearly showed that virus culture can be established from dry swabs stored for days at RT in TE buffer devoid of proteinase K (Ram et al., 2021). Shortly, the dry swab method conserves all the advantages of gold-standard method and can deliver reliable results even faster. In the present scenario, implementation of this method could provide multiple advantages in containing the infection and better allocation of medical resources in the developed and developing nations alike.

## Data Availability

All the data related to this study are provided in the main text or in the supplemental files.

## Ethical Statement

The study follows the institutional ethics committee guidelines. Informed consents were taken from the participants of the optimisation of proteinase K concentration study.

## Acknowledgements

UK and CGG thank the financial support received from UGC and CSIR, India, respectively. All the authors acknowledge the support received from CSIR, India. The authors gratefully acknowledge Divya Gupta, Vishal Sah, and Krishnan H Harshan, CSIR-CCMB for providing the cultured viral strains.

