## Supplementary Figure for "Temporal stability and detection sensitivity of the dry swab-based diagnosis of SARS-CoV-2"

**Supplementary Figure 1**

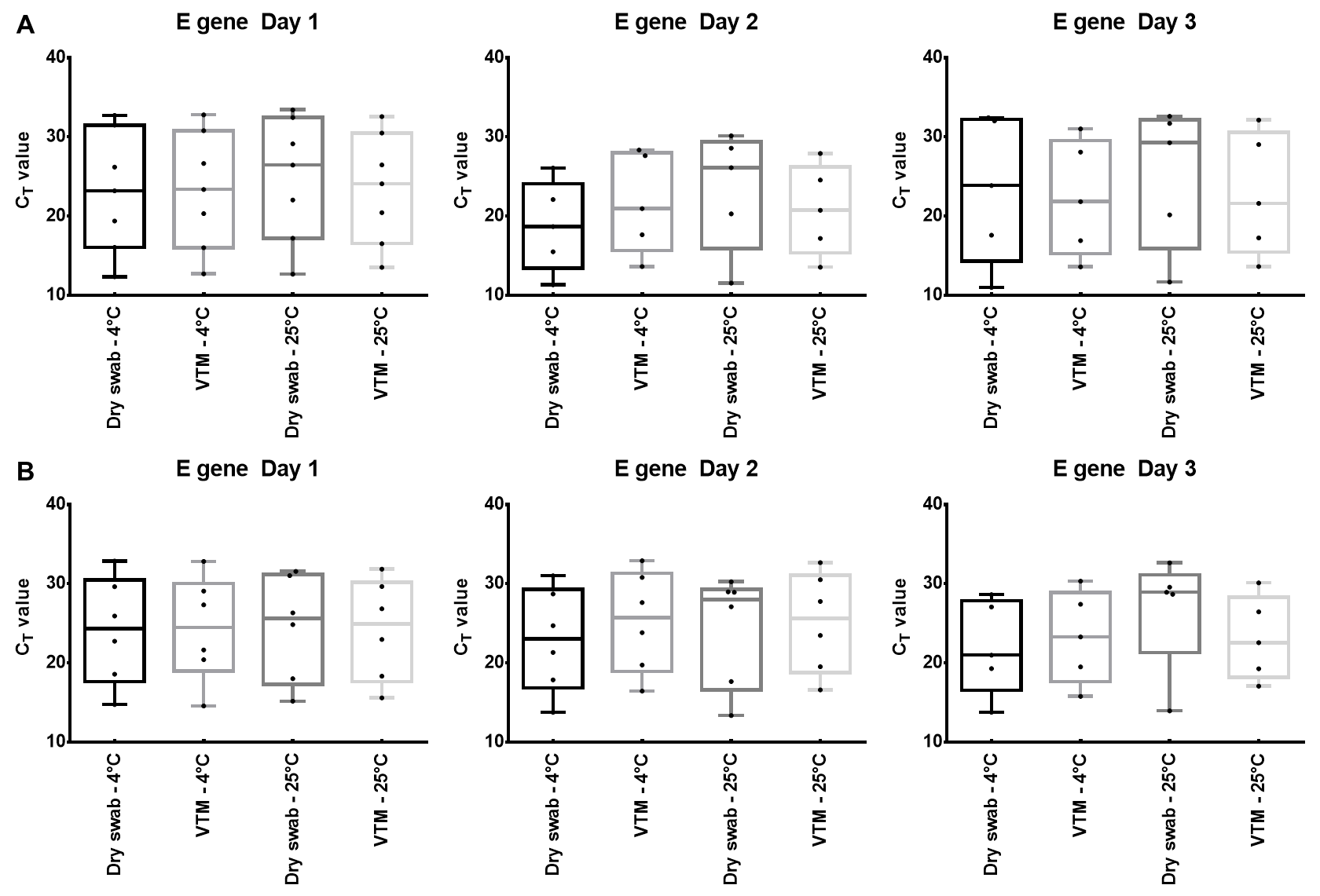

**Supplementary Figure 1**: Comparison of E gene CT values of the samples stored in different conditions for the mentioned time. The data indicates no loss in detection when dry swabs are stored at RT for 1 day and up to 3 days when stored at 4℃. Data shown are from SARS-COV-2 (A) A2a strain and (B) A3i strain. RT-PCR was performed in triplicates for each sample and the average CT values are plotted.

**Supplementary Figure 2**

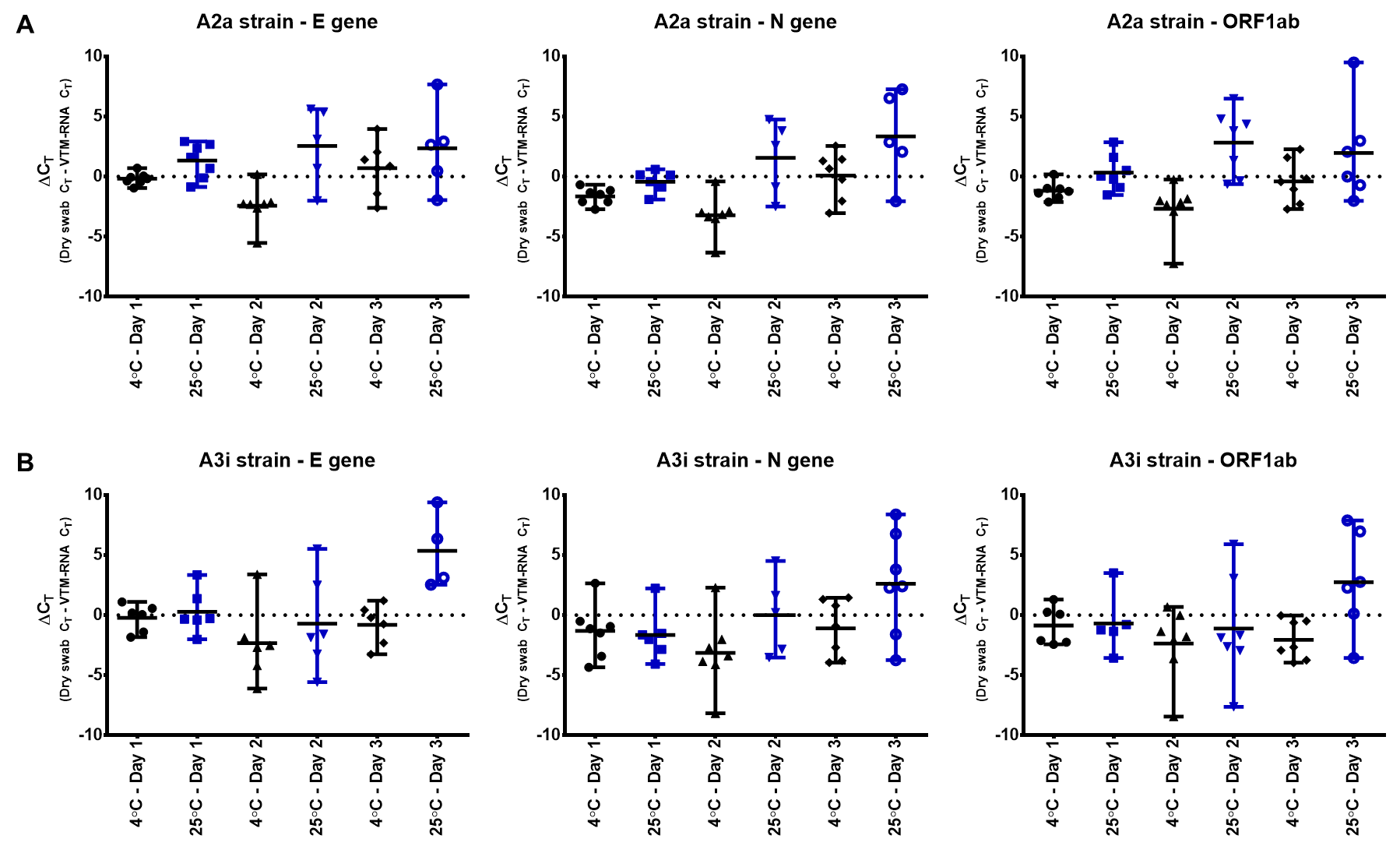
**Supplementary Figure 2:** Graphs indicating the distribution of the delta C_T_ values of the target genes from samples subjected to various conditions - (A) A2a strain; (B) A3i strain. The delta C_T_ values were calculated by subtracting the dry swab samples’ C_T_ values from that of the VTM-RNA samples. The horizontal bar indicates the mean delta C_T_ values. Black coloured vertical bars shows the range of the C_T_ values of samples stored at 4℃ while the blue vertical bars represents C_T_ values of samples stored at RT.
