## Supplementary Table for "Temporal stability and detection sensitivity of the dry swab-based diagnosis of SARS-CoV-2"

**Supplementary Table 1: Correlation coefficients of the C_T_ values of dry swab samples versus VTM-RNA samples.**

| **A2a** | **Day-1** | | **Day-2** | | **Day-3** | |
| --- | --- | --- | --- | --- | --- | --- |
|  | **4℃** | **25℃** | **4℃** | **25℃** | **4℃** | **25℃** |
| E gene | 0.998 | 0.986 | 0.975 | 0.920 | 0.975 | 0.919 |
| N gene | 0.996 | 0.995 | 0.973 | 0.908 | 0.982 | 0.951 |
| ORF1ab | 0.997 | 0.979 | 0.958 | 0.950 | 0.974 | 0.879 |
| **A3i** | **Day-1** | | **Day-2** | | **Day-3** | |
|  | **4℃** | **25℃** | **4℃** | **25℃** | **4℃** | **25℃** |
| E gene | 0.986 | 0.961 | 0.879 | 0.825 | 0.975 | 0.771 |
| N gene | 0.955 | 0.947 | 0.882 | 0.944 | 0.952 | 0.781 |
| ORF1ab | 0.971 | 0.904 | 0.896 | 0.798 | 0.972 | 0.804 |

**Supplementary Table 2: Correlation coefficients of the C_T_ values of dry swab samples stored for 3 days vs 1 day.**

| **Dry swabs** | **A2a** | | **A3i** | |
| --- | --- | --- | --- | --- |
|  | **4℃** | **25℃** | **4℃** | **25℃** |
| E gene | 0.956 | 0.927 | 0.962 | 0.799 |
| N gene | 0.964 | 0.974 | 0.941 | 0.768 |
| ORF1ab | 0.950 | 0.900 | 0.961 | 0.823 |

|  |  | **Nasopharyngeal samples** | | |
| --- | --- | --- | --- | --- |
| **Sample** | **ProK Conc. (mg/ml)** | **E gene Ct** | **N gene Ct** | **ORF1ab Ct** |
| **Sample 1** | **2 mg/ml** | 18.50 | 16.50 | 18.10 |
|  | **1 mg/ml** | 17.50 | 15.37 | 15.86 |
|  | **0.5 mg/ml** | 17.40 | 15.41 | 15.59 |
|  | **0 mg/ml** | 20.19 | 17.01 | 17.50 |
| **Sample 2** | **2 mg/ml** | 31.00 | 29.80 | 30.70 |
|  | **1 mg/ml** | 31.05 | 28.55 | 29.27 |
|  | **0.5 mg/ml** | 30.81 | 27.96 | 28.94 |
|  | **0 mg/ml** | 31.13 | 28.11 | 29.45 |
| **Sample 3** | **2 mg/ml** | 12.20 | 10.50 | 12.40 |
|  | **1 mg/ml** | 10.78 | 10.12 | 10.63 |
|  | **0.5 mg/ml** | 8.83 | 7.31 | 7.30 |
|  | **0 mg/ml** | 12.27 | 9.57 | 10.02 |

**Supplementary Table 3: C_T_ values of the samples treated with different proteinase K concentrations.**
